# Identifying biomedical entities for datasets in scientific articles – A 4-step cache-augmented generation approach using GPT-4o and PubTator 3.0

**DOI:** 10.1101/2025.03.04.25323310

**Authors:** Claudia Giuliani, Gita Benadi, Felix Engel, Jonas Werner, Manuel Watter, Guido Schwarzer, Olaf Groß, Robert Zeiser, Harald Binder, Klaus Kaier

## Abstract

The accurate annotation of biomedical entities in scientific articles is essential for effective metadata generation, ensuring data findability, accessibility, interoperability and reusability in collaborative research. This study introduces a novel 4-step Cache-Augmented Generation (CAG) approach to identify biomedical entities, leveraging GPT-4o and PubTator 3.0. The method integrates (1) GPT-4o-based entity generation, (2) PubTator-based validation, (3) term extraction based on a metadata-schema developed for the specific research area, and (4) a combined evaluation of PubTator-validated and schema-related terms. Applied to 23 articles published in the context of the Collaborative Research Centre *OncoEscape*, the process was validated through supervised, face-to-face interviews with article authors, allowing an assessment of annotation precision using random effects meta-analysis. The approach yielded a mean number of 19.6 schema-related and 6.7 PubTator-validated biomedical entities per article. Overall precision was 98% [95%CI 94%-100%]. In a subsample (N=20), available supplemental material was included in the prediction process, which did not increase precision (98%, CI 95%-100%). Moreover, the mean number of schema-related (20.1, p=0.561) and PubTator-validated (6.7, p=0.681) biomedical entities did not increase with the additional information provided with the supplement. This study highlights the potential of CAG for metadata annotation. The findings underscore the practical feasibility of full-text analysis for routine metadata annotation in biomedical research.

## Introduction

Long-term university-based research institutions, such as the German Collaborative Research Centres (CRCs) which can be funded for up to 12 years, require controlled and structured sharing of data and documents, with accurate, interoperable metadata being crucial for long-term data reusability and understanding [2, 3] in accordance with the FAIR principles [4]. For biomedical research data, a particularly important type of metadata are biomedical entities such as organisms, cell lines or genes. If published research articles and the associated datasets are tagged with these biomedical entities, it is easier for scientists to find relevant datasets for their research and thus reuse these data. Since manual entity annotation is often a time-consuming task, it is desirable to automate this process as much as possible. One method of automatic biomedical entity identification is named entity recognition, which involves identifying spans of text that represent named entities (e.g. “mouse”) and tagging them with the appropriate category (e.g. “organism”) [5]. Entity recognition is challenging due to difficulties in segmentation, i.e., determining whether (1) a term represents an existing biomedical entity that is (2) appropriate for describing the datasets that are presented in the respective research article.

Large Language Models (LLMs) can be used for entity recognition and they can significantly increase the speed of biomedical entity documentation [6]. While LLMs exhibit strong language capabilities, they are prone to generating incorrect information, often referred to as hallucinations [7, 8]. A possible way of mitigating this problem is to restrict the terms identified by the LLM to a set of known true bio-entities. This restriction can either be directly specified in the prompt to the LLM, or the list of terms suggested by the LLM can be validated in retrospect.

In this manuscript, we present a newly developed method for article-based metadata annotation prediction based on the usage of LLMs and the Cache-Augmented Generation (CAG) method [11]. In particular we used the LLM GPT-4o in combination with PubTator 3.0, a tool built by the National Library of Medicine to identify several entities in the literature using state-of-the-art Artificial Intelligence (AI) techniques [9, 10], and a dedicated metadata-schema developed to describe data collected for a specific research area [12]. With this twofold combination of validation and constraint of the LLM we intend to provide a reliable and extended list of annotation suggestions exploiting the strengths of both PubTator, like comprehensive genes and chemicals lists, and of the dedicated metadata schema, like relevant mouse and cell lines which are often created in-house. The approach consists of four steps: (1) GPT-4o-based full-text analysis and suggestion of biomedical terms; (2) PubTator-based validation of suggested terms from step one; (3) Full-text analysis restricted to schema-related terms; (4) Combination of schema-related and PubTator-validated terms.

We applied this 4-step approach in the context of the CRC *OncoEscape* (CRC 1479) and tested the feasibility of the approach using a structured, paper-based, supervised annotation double-checking process. This process involved face-to-face interviews with the scientists responsible for authoring the respective article and enabled us to calculate a first estimate of the proportion of correctly predicted annotations.

## Methods

We included a total of 23 articles that were published during the first funding period of *OncoEscape* (2021-2025), and for which a scientist responsible for authoring the article had agreed to take part in face-to-face interviews (Table S1 in supplemental material). We deliberately accepted this restriction, as we assume that the social control arising from the presence of a member of the research data management group is most appropriate to obtain a reliable assessment of the precision of the 4-step approach. All participants in the face-to-face interviews carefully checked the annotation predictions. In a subsample (N=20), available supplemental material was also included in the prediction process and two prediction results were validated in the face-to-face interviews. Of the three remaining articles, one had no supplemental material, one had such a large supplemental-material section that the LLM was not able to analyze it correctly, the third was not considered in the face-to-face interviews.

### LLM-based annotation prediction

A systematic 4-step approach was used for annotation prediction (**Figure 1**): (1) ChatGPT was initially tasked with generating relevant entities based on the paper full text. No further restrictions or suggestions were made but the LLM was instructed to ignore the discussion and bibliography for its predictions. (2) The resulting entities were then validated using the PubTator 3.0 database. (3) Next, ChatGPT was instructed to reanalyze the full text of the manuscript but with the task to identify entities defined in a predefined schema. For this scope, a previously developed dedicated *OncoEscape* metadata schema [12] was included in this prompt in a tree-like structure. (4) Finally, a combined evaluation of the results from steps 2 and 3 was prompted, with the clear order of listing only those PubTator-validated entities that had not already been identified through the schema extraction in step 3.

**Figure 1:**
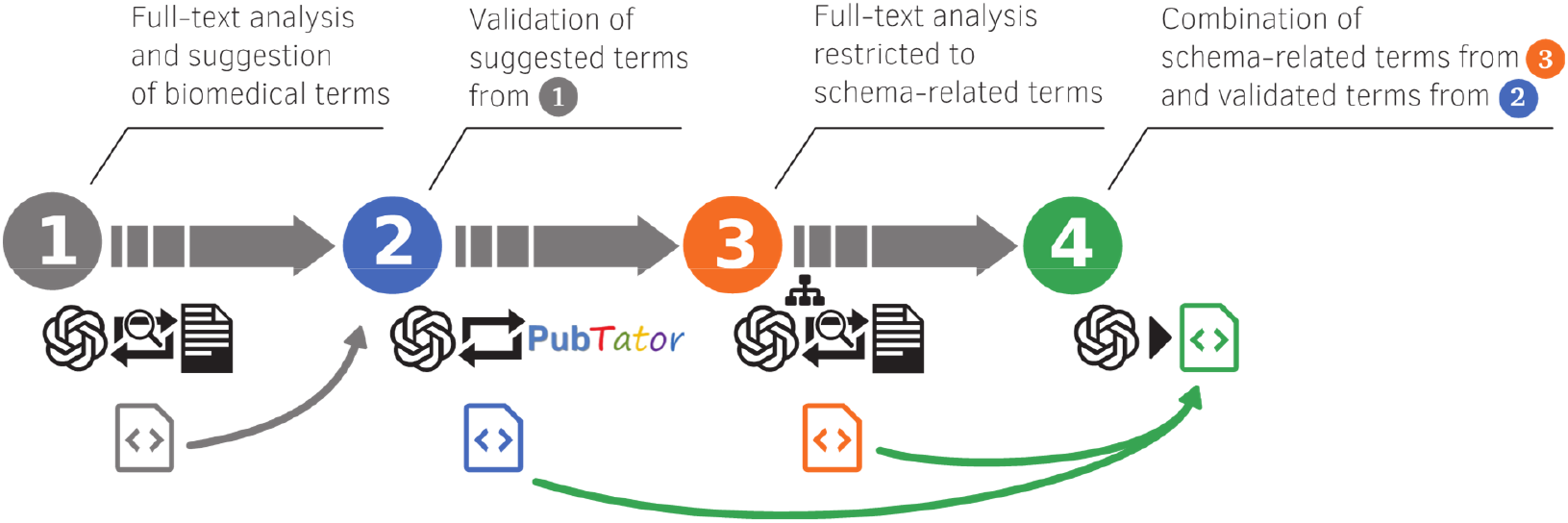
4-step cache-augmented generation approach for annotation prediction

This process requires comprehensive textual analysis and the integration of knowledge, a method commonly referred to as Cache-Augmented Generation (CAG). In recent years, Retrieval-Augmented Generation (RAG) has emerged as the standard approach for customizing large language models (LLMs) to meet specific informational needs. However, with recent advancements in long-context LLMs, it is now feasible to eliminate the need for RAG by directly embedding all proprietary information within the prompt [11]. RAG’s reliance on real-time retrieval could introduce latency and potential errors in document selection, especially with large datasets. CAG, by preloading all relevant resources into the LLM’s extended context, has been found to eliminate retrieval latency and minimize errors.

Based on the recommendations of the developers of PubTator 3.0 [10], we created a custom GPT with PubTator 3.0 augmentation following the instructions of the authors [13]. The above described 4-step approach was operationalized using separate prompts in the chat interface from OpenAI, where the publication text was uploaded in the first step as a PDF. See supplemental material for further details of the 4-step approach and specifically Table S2 for the used prompts. All analyses were conducted using ChatGPT-4o-2024-11-20. Please note that this multi-step prompting approach involved an iterative process in which ChatGPT connected to PubTator 3.0 to ensure reliability of the results. For each entity generated in step (1), step (2) instructs ChatGPT to query PubTator in order to retrieve a standardized entity ID, ensuring the validation of suggestions and alignment with recognized biomedical terms. Step (3) serves the purpose of aligning and categorising the biomedical entities suggestion with the dedicated *OncoEscape* metadata schema. This iterative process of generation, validation, and evidence gathering ensures that only scientifically grounded and relevant entities are carried forward and thereby overcomes some of the hallucination problems arising when prompting ChatGPT in an unspecific way. Furthermore, the multi-step prompting approach enables us to separate the capabilities of annotation prediction based on a schema and those of PubTator. In other words, the Pubtator-based predictions can consider the scientific context from the paper and are not limited by the schema incompleteness. The PubTator-validated entities are either associated to a category, but were not included in the list of schema-predefined entities, or are entities which could not be associated to any *OncoEscape* schema category. Whereas PubTator has six categories of annotated entity types, Gene, Disease, Chemical, Variant, Species and Cell Line, the “FindEntityID” endpoint used in the GPT+PubTator implementation does not consider Species and Cell Line entries. This limitation is not relevant for the overall suggestion completeness since the OncoEscape metadata schema contains relevant species and dedicated cell lines used in the specific research area.

### Validation of LLM-based annotation prediction

In order to measure the prediction capabilities of the LLMs, precision of prediction is measured on the level of a single study as a proportion of correctly predicted annotations. To identify correctly predicted annotations, we employed a supervised annotation double-checking process. This process involved face-to-face interviews between a member of the Research Data Management (RDM) group and a senior scientist responsible for authoring the article. Scientists were instructed to consider the datasets referenced in the article as the context for evaluating annotations. Each annotation suggested by the 4-step approach was examined to determine whether it accurately described the datasets used for the article. If an annotation was suspected to be incorrect, participants were encouraged to consult the methods or results sections of the article, which was provided in paper for quick verification. Annotations deemed incorrect based on suspicion or evident errors could also be marked incorrect without comprehensive cross-checking to prioritize efficiency. The process was conducted in a supervised manner, and a five-minute time span was recommended for a single article. This five-minute face-to-face evaluation method ensured a practical balance between efficiency and reliability, enabling the rapid assessment of the 4-step approach.

### Statistical analysis of the interview outcomes

To estimate the LLM prediction accuracy across all studies, methods for the meta-analysis of single proportions are applied [14]. Due to the small number of entities per study, the Freeman-Tukey double arcsine transformation was used [15]. This transformation stabilizes the variances, makes the meta-analysis more robust when proportions are close to 1 and enables working with small sample sizes. A random-effects model using the REML estimator for the between-study variance was considered. Also, the behaviour of the LLM model, which is intrinsically stochastic, as well as the possible variability in the evaluation of LLM-suggestions by different authors justify the calculation of confidence intervals for the accuracy predictions in individual papers. Differences between the number of article-based and article+supplement-based biomedical entities were analysed using the Wilcoxon signed-rank test.

## Results

A total of 604 biomedical entities were predicted for the 23 articles (Figure 2). This equals a mean number of 26.3 biomedical entities per article of which 19.6 were schema-related and another 6.7 were PubTator-validated biomedical entities. Overall precision, defined as ratio of correct entities over total number of annotation suggestions, was 98% [95%CI 94%-100%], meaning that the vast majority of predicted biomedical entities were considered correct in the face-to-face interviews.

**Figure 2:**
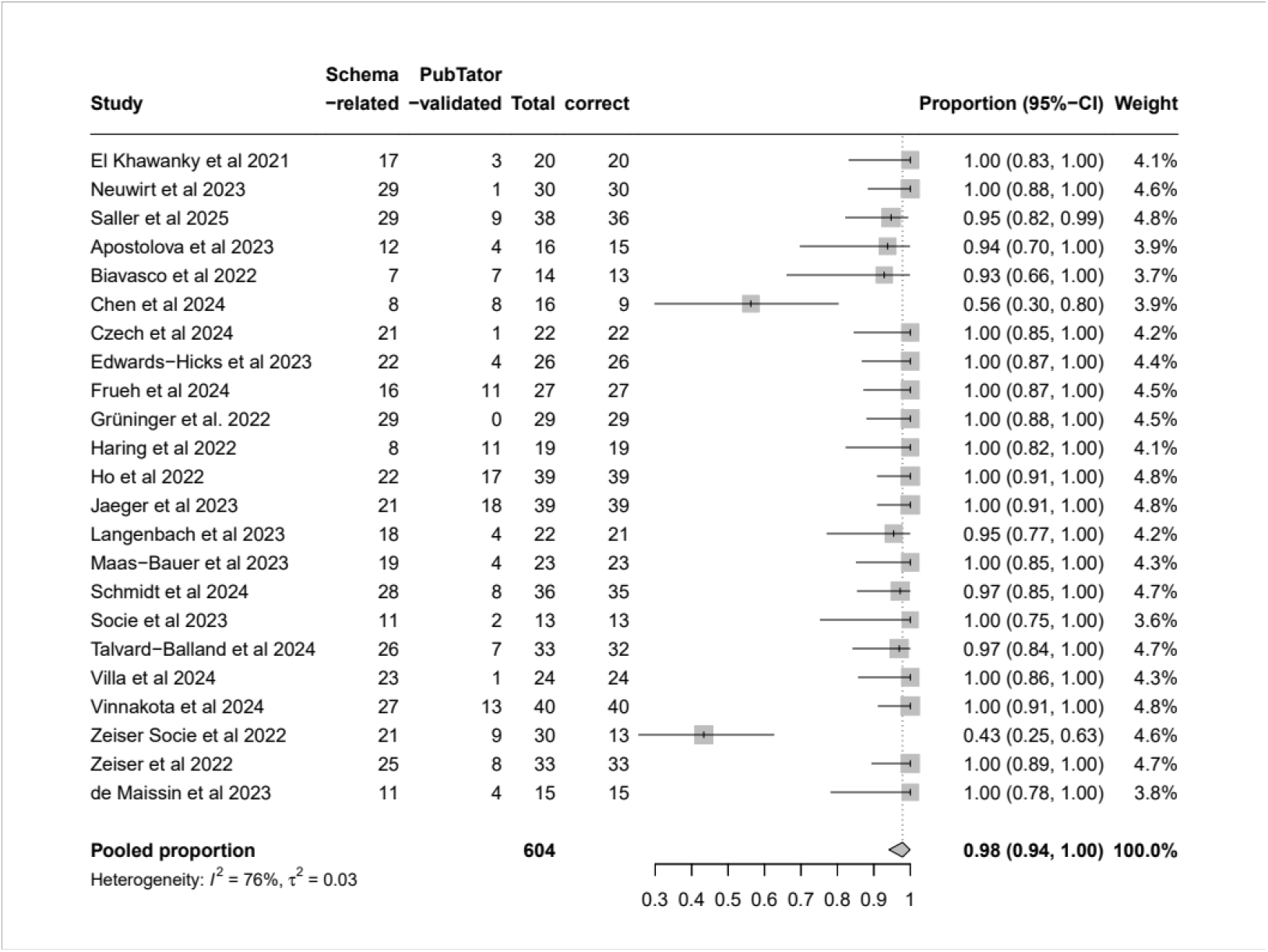
Precision of annotation predictions for each paper without supplement considered in this study together with pooled precision, heterogeneity and τ^2^.

Interestingly, the precision of schema-related biomedical entities was 97% [CI 91%-100%], while the precision of PubTator-validated biomedical entities was 100% (supplemental Figure S1 and S2). Most of the articles had a proportion of correctly chosen entities for the annotation close or equal to 100% (Figure 2). Only two papers, Chen et al 2024 and Zeiser Socie et al 2022, had a markedly lower proportion of correctly suggested annotation entities which contributed to the considerable between-study heterogeneity (I^2^ = 76%). Both papers are clinical studies concerned with the transfer of basic research results into a clinical setting and had a number of false-positive basic research-related annotations that, although mentioned in the text, served as a rationale for the clinical approach rather than describing the underlying dataset. A post-hoc sensitivity analysis excluding these two studies resulted in a pooled precision of 100% [99%-100%] without important between-study heterogeneity (I^2^ = 0%).

In a subsample (N=20), available supplemental material was also included in the prediction process and two prediction results, both those for the paper with and without supplement, were validated in the face-to-face interviews (see Figure S3 in the supplemental material). Interestingly, availability of supplemental material in the prediction process did not increase precision (98%, CI 95%-100%) with between-study heterogeneity I^2^ = 67%. Moreover, the mean number of schema-related (20.1, p=0.561) and PubTator-validated (6.7, p=0.681) biomedical entities did not increase with the additional information provided with the supplement and are compatible with the values obtained with the paper-only approach.

No statistically relevant differences were observed in the number of suggested entities for the papers without or with supplement also when considering the annotation suggestions per category (Figure 3). Here only the 20 papers including annotation suggestions for the supplement are considered. The listed categories are as defined in the *OncoEscape* schema with two additional categories representing the availability of data, DataAvailability, which can be available publicly or on request as extracted from the paper, and the PubTator-related entities that the LLM could not assign to a predefined category, PubTatorNotClassified. Additional distributions are shown in the supplemental material.

**Figure 3:**
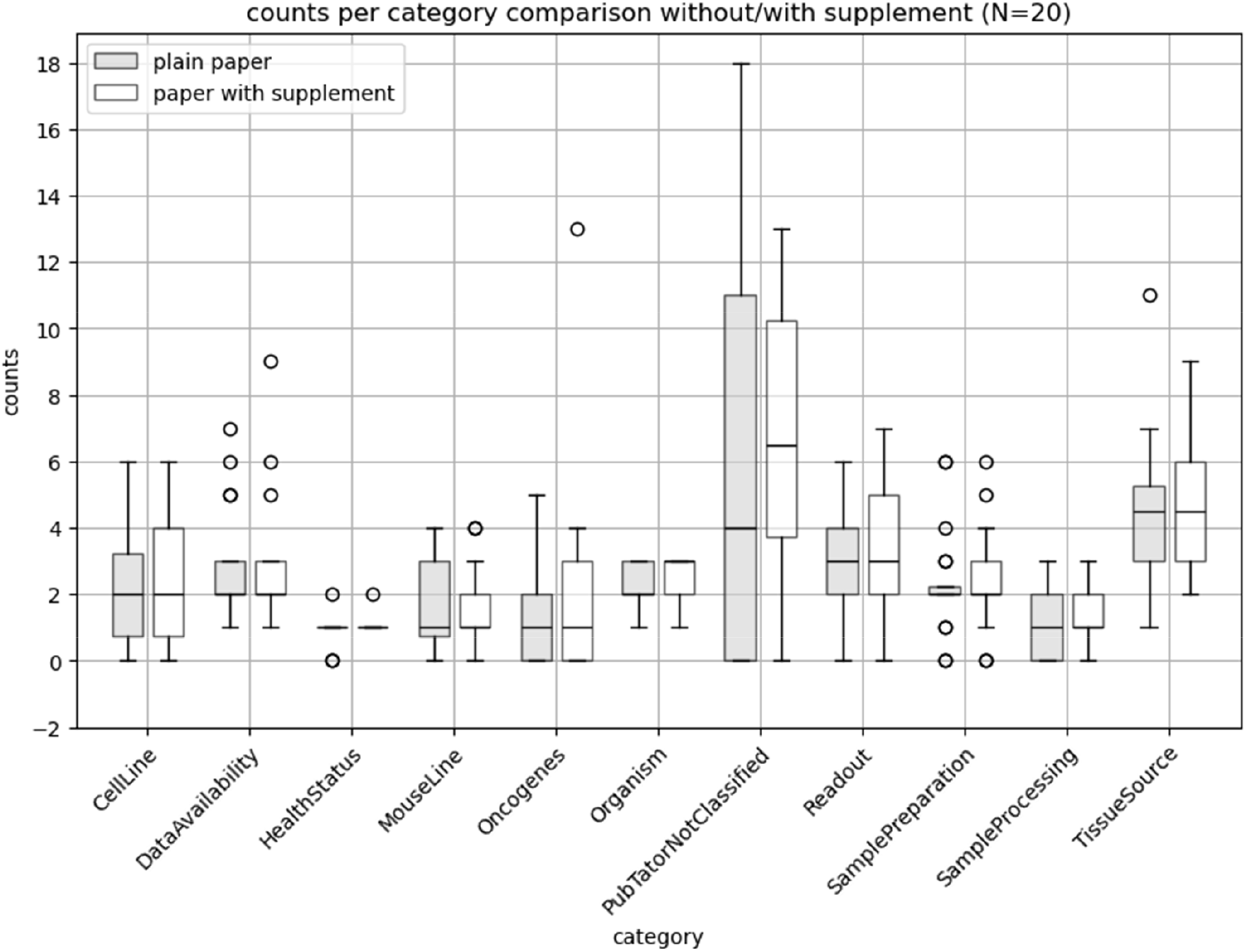
Counts of LLM annotation suggestions per category for the paper without and with supplement. Only papers with an annotated supplement are considered (N=20).

The compatibility of the annotation suggestions obtained by analyzing the paper with or without the supplement is confirmed, both for the overall number of suggestions (p=0.556) as well as for the suggestions divided per category. The detailed results of the Wilcoxon signed-rank-tests are summarized in Table S3 of the supplemental material.

## Discussion

Our 4-step approach involves a combination of GPT-4o, PubTator, and schema-related term analysis. This process requires comprehensive textual analysis and the integration of knowledge in an iterative process. We exploit CAG by preloading all relevant resources into the LLM’s extended context to accelerate information retrieval and minimize response errors. For instance, since both steps 1 and 3 involve full-text analysis, we can accelerate step 3 by using CAG. Preloading the cache allows the model to quickly access and process the entire article text, leading to a fast suggestion of terms [11]. Moreover, CAG ensures that the model has a unified understanding of the entire article context, leading to improved consistency and accuracy in term prediction across all steps. This is crucial for maintaining the coherence and quality of metadata annotation throughout the process.

The high precision observed in our study underscores the potential of the CAG approach for streamlining metadata annotation. Nevertheless, problems with very long articles or extensive supplemental materials can arise when the available LLM context length is exhausted. Recent studies have shown that while LLMs benefit from larger context windows, their performance does not always scale linearly with increased input size, often exhibiting a “lost in the middle” effect where information in the middle of long texts is processed less effectively [16]. Further studies about the effect of the LLM-model choice and its context length on the reliability of the annotation predictions are ongoing. Moreover, since the supplemental material did not improve the average number of annotation suggestions or the proportion of correct ones, and since it is complicated to retrieve the supplemental material in a standardized way, we deem that the LLM annotation suggestions of papers without supplement should suffice in most cases.

It is to be noted that the provided metadata schema has a non-negligible impact on the quality and level of details of the biomedical entity suggestions and categorization by the LLM, especially for the categories not covered by the PubTator entities, e.g. readout, sample preparation and processing, tissue source. For example, if a dataset was processed with single cell RNA-sequencing but we give the LLM only first level of details of readout methods, it will then be able to provide only the given level of detail. In this case “Sequencing” will be selected as annotation suggestion, which could be deemed wrong by the authors of the paper. On the other hand, if we provide the LLM with additional readout levels of detail and, continuing with our example, we include e.g. different sequencing options, the LLM is able to identify that the data were processed with “single cell RNA-sequencing”. Moreover, if the schema entities are passed to the LLM in a tree-like structure, the LLM is able to identify dependencies and to reproduce the annotation depth. Likewise, the schema limits the capabilities of the LLM to identify correctly entities, especially if the paper in question does not analyze primary datasets in the context of basic research, but is e.g. a clinical or modeling study. This is apparent in the lower proportion of correctly suggested entities in the papers discussing clinical studies. In both such articles the basic-research rationale was applied for the clinical approach, but the underlying datasets were of clinical nature.

In general, the LLM annotation should allow to have a great number of papers and datasets annotated, which increases the FAIRness of the data. A comprehensive assessment not only of the quantitative, but also of the qualitative advantages of the LLM automatic-annotation versus a diligent manual annotation is being investigated. It is already apparent that including the LLM suggestions and their validation through PubTator is beneficial for having a comprehensive list of (onco)genes and treatments considered in the experiments underlying the datasets. The PubTator-validated entities identified outside the schema highlight the potential for a hybrid approach that balances structured annotation with the flexibility to capture emerging concepts. Future work should explore dynamic schema adaptation, e.g. to include other areas of research such as computational modeling, and integration of knowledge graphs [17, 18] to represent complex relationships between biomedical entities, ultimately enhancing the comprehensiveness and accuracy of metadata annotation.

A major challenge of our approach was the distinction between article and datasets: the metadata annotation was focussed on the respective underlying datasets, however, the prediction was conducted for each already published journal publication. This was intended, as we assume that scientists can usually refer to the respective article when annotating their datasets with metadata. However, an article does not only describe details of the respective datasets, but also discusses aspects that go beyond them. We tried to solve this discrepancy by instructing the LLM to ignore the discussion and bibliography of the full texts during prediction (step 1). In view of some incorrect predictions, however, this may not have worked entirely. Nevertheless, we felt it was extremely important to further develop our 4-step approach on the entire full text of the article (instead of just feeding in parts of the article), as this seems to be the only practicable way to transfer it into a fully automated annotation process.

Our aim is to enrich the datasets collected within the research projects with metadata information to ultimately make them FAIR. To lessen the problem of having annotation predictions extracted from the analysis of the article and being unable to assign them reliably to a given dataset, we let the LLM retrieve information about available datasets after running the 4-step approach. For published datasets, the LLM was able to find the links to the datasets in the article and, when asked for it, could directly gather information about each published dataset through a web search. At least for published datasets we can thus reliably assign our annotation suggestions to a specific dataset. However, some datasets are not published or only available on request. Since in any cases some datasets are not fully described, we can only say that data with certain properties and preparation were collected, but we cannot have a 1 to 1 assignment of metadata to datasets. To improve the FAIRness of these latter datasets additional infrastructural or institutional measures need to be taken in combination with the further development of the LLM analysis methods. Additional measures could be the standardization of the datasets format, inter-connection of systems for automatic collection of metadata during the complete data lifecycle (data storage solutions, electronic lab notebooks, data collection machines, e.g. microscopes), or requirements that all dataset should be eventually published.

In studies on NLP using LLMs, false negatives are often also considered to determine the accuracy, recall and the F1 score. In the present situation, where the goal of prediction is to maximize the number of correct annotations while also keeping the proportion of false positive annotations as small as possible, this is not possible because the number of potential annotations per dataset has no clearly defined upper limit. Although it would be possible to ask the authors in the face-to-face interviews which biometric entities they are missing, this question cannot be answered without further effort. Moreover, we actively constraint the LLM by giving it information about the predefined CRC metadata schema so that we not only have sensible annotations depending on the paper context, but more importantly also relevant ones in connection to the CRC.

Finally, further studies to evaluate the reproducibility of results given the stochastic nature of the LLM output and the variability of human responses in interviews need to be done. In particular we plan to investigate the impact of the choice of LLM model on the annotation suggestions, as well as the variation of number of suggestions when running the 4-step-approach multiple times on a single paper.

## Data Availability

All data produced in the present study are available upon reasonable request to the authors

