## Supplemental Material for "Identifying biomedical entities for datasets in scientific articles – A 4-step cache-augmented generation approach using GPT-4o and PubTator 3.0"

*Table S1: List of OncoEscape Papers, naming as in Paper, used for the evaluation of the 4-step cache-augmented generation approach using GPT-4o and Pubtator 3.0 and their PubMedIDs*

| Name | PubMedID |
|---|---|
| El Khawanky et al. 2021 | 34750374 |
| Neuwirt et al. 2023 | 36649377 |
| Saller et al. 2025 | 39571574 |
| Apostolova et al 2023 | 37539479 |
| Biavasco et al 2022 | 35768570 |
| Chen et al 2024 | 38844797 |
| Czech et al 2024 | 38381845 |
| Edwards-Hicks et al 2023 | 36732424 |
| Frueh et al 2024 | 38170159 |
| Grüninger et al. 2022 | 35794338 |
| Haring et al 2022 | 34407601 |
| Ho et al 2022 | 35853161 |
| Jaeger et al 2023 | 36696631 |
| Langenbach et al 2023 | 37071397 |
| Maas-Bauer et al 2023 | 38199985 |
| Schmidt et al 2024 | 38885318 |
| Socie et al 2023 | 36827620 |
| Talvard-Balland et al 2024 | 38916965 |
| Villa et al 2024 | 38200005 |
| Vinnakota et al 2024 | 38741011 |
| Zeiser Socie et al 2022 | 34971577 |
| Zeiser et al 2022 | 35081254 |
| de Maissin et al 2023 | 37439488 |

**Prompt Strategy**

The prompts used for the 4-step strategy are added below (Table S2).

For machine readability of the results and to collect information on the availability of the datasets, three additional queries were run: 1) Dataset availability (public or on request); 2) If public available datasets exist, information about them should be retrieved, if possible with the same categorisation as for the paper; 3) consolidated results, with data availability, in tabular form for machine readability and analysis of results.

*Table S2: Prompt used for the 4-steps approach.*

| Step | Prompt |
|---|---|
| 1 | **Identification of Biomedical Entities**<br>Analyze the uploaded manuscript to identify all biomedical entities mentioned. Exclude entities mentioned only in the discussion section or the bibliography. Provide a |

| | |
|---|---|
| | comprehensive list of these entities. Complete this step entirely before proceeding to the next step. |
| 2 | **PubTator Analysis**<br>For each biomedical entity identified in Step 1, determine whether it is listed in PubTator. Provide a list of these entities along with their existence status in PubTator (e.g., listed or not listed). Do not include examples or elaborate explanations. Complete this step entirely before proceeding to the next step. |
| 3 | **Manuscript Reanalysis for Specific Aspects**<br>Reanalyze the uploaded manuscript to identify the presence of specific aspects listed below under each subheading. Only include aspects explicitly mentioned in the manuscript, excluding mentions from the discussion section or bibliography. Maintain the formatting and organization provided below, and provide a comprehensive list for each subheading.<br>**Organism**<br>• Cell line<br>• Human<br>• Mouse<br>**List of Cell Lines**<br>• B16.F10<br>• B16.F10$^{luc/GFP}$<br>• B16.F10$^{OVA}$<br>• AT-3$^{OVA}$<br>• RENCA<br>• KP1.9<br>• 4434-BRAF$^{V600E}$<br>• MC38<br>• MC38$^{OVA}$<br>• MC38$^{ROR1+/GFP+/Luc+}$<br>• RMB1<br>• C1498<br>• C1498$^{GFP+ luc+}$<br>• Yumm1.7<br>• Yumm1.7$^{OVA}$<br>• MODE-K<br>• JIMT-1 breast cancer<br>• 32D<br>• Platinum E<br>• WEHI-3B<br>• WEHI-3B$^{Luc/GFP}$<br>• WEHI-3B$^{CD155KO}$<br>• MOLM-13<br>• BAF-3<br>• MV4-11<br>• MV4-11$^{Luc+}$<br>• MV4-11$^{NT Luc+}$<br>• MV4-11$^{JunD KO (G4) Luc+}$<br>• MV4-11$^{cJun KO (G11) Luc+}$<br>• MV4-11$^{JunD KO (G64) Luc+}$<br>• MV4-11$^{cJun KO (G69) Luc+}$<br>• MV4-11$^{AXL KO (G1) Luc+}$<br>• OCI-AML2$^{GFP+Luc+}$<br>• OCI-AML3<br>• OCI-AML3$^{GFP+Luc+}$<br>• Kasumi-1 |

- THP-1
- HL-60
- SEM
- E2a-PBX
- A20$^{\text{GFP+Luc+}}$
- OCI-AML3$^{\text{hp53 1961 (p53 KD) YFP+ dsRed+}}$
- OCI-AML3$^{\text{hp53 Renilla (p53 WT) YFP+ dsRed+}}$
- RP1199.6
- RP1199.1
- RP1201.1
- RP1209.1
- RP12086
- Panc$^{\text{ROR1+/GFP+/Luc+}}$
- STC-1
- NB-4
- ML-2
- K562
- KG-1
- KG-1α
- MUTZ-8
- BV2
- FL83B
- HLE
- Colo800
- murine embryonic fibroblasts
- OV-90
- BxPC3
- NCI-H2405
- HEK293T
- Steinberger NFAT-reporter
- SMMC-7721
- HepG2

**Tissue Source**

- Adrenal gland
- Blood
    - Blood plasma
    - Blood serum
    - Whole blood
- Bone marrow
- Brain
- Embryonal tissue
- Heart
- Intestine
- Kidney
- Liver
- Lung
- Lymph node
- Nerve
- Skin
- Spleen
- Thymus
- Urine

- Feces
- Vascular system

**Health Status**

- Cancer

**Mouse Line**

- C57BL/6J (wildtype)
- C57BL/6J (wildtype)
- C57BL/6-Nrastm1Tyj/J X Vav-Cre
- C57BL/6JCya-Tigitem1/Cya
- Tigitfl/fl;CD4cre/+
- CD155$^{-/-}$ mice
- Tigitfl/fl;Zbtb46cre/+
- Gal9$^{-/-}$
- Nlrp3$^{-/-}$
- Pycard$^{-/-}$
- Casp1$^{-/-}$
- Nt5e/Cd73$^{-/-}$
- Gsdmd$^{-/-}$
- Il1r1$^{-/-}$
- Cmtm6$^{-/-}$
- Osm$^{-/-}$
- Osmr$^{-/-}$
- FLT3-ITD
- Ddit3$^{-/-}$
- Rag1$^{tm1Mom}$
- Rag2$^{-/-}$gc$^{-/-}$
- Tet2-ko (B6(Cg)-Tet2tm1.2Rao/J
- Apcflox/flox
- Col7a1$^{fl/fl}$
- Pycard $^{fl/fl}$
- Xbp1 $^{fl/fl}$
- Adora2 $^{fl/fl}$
- Dnmt3afl/+
- Osmr $^{fl/fl}$
- Il1r1 $^{fl/fl}$
- Trp53fl/fl
- Trp53LSL-R175H
- KrasLSL-G12D
- Vhl $^{fl/fl}$
- Rb1 $^{fl/fl}$
- Tak1 $^{fl/fl}$
- Atf6 $^{fl/fl}$
- Ptpn11 $^{D61Y/+}$
- SCLtTA/TRE-Cre
- SCL-Cre
- Osterix-Cre
- LepR-Cre
- Prx1-Cre
- CD4-Cre
- CX3CR1 ERT2 Cre
- HexbCreERT2:R26R$^{Confetti}$
- Mrc1CreERT2:R26R$^{Confetti}$
- Cxcr4CreERT2:R26R$^{Confetti}$

- Ksp1.3-Cre$^{ERT2}$
- Ksp1.3-Cre$^{ERT2}$; Vhl $^{fl/fl}$; Trp53 $^{fl/fl}$; Rb1 $^{fl/fl}$
- Mx1 Cre
- LysM Cre iDTR
- Cd4 Cre ERT2
- Villin-CreERT2
- Rosa26::CreERT2
- Tet2 $^{fl/fl}$Mx1-Cre
- Rosa26::Cre ERT2 Kras$^{G12V}$
- Rosa26::Cre ERT2 Jak2-V617F FLEX/+
- nATF6liv
- Cas9
- MMTV-PyMT
- ERAI
- OT-1/Rag2$^{-/-}$

**Sample Preparation**

- Cultured cells
  - Adipocyte
  - Cardiomyocyte
  - Dendritic cell
  - Embryonic cardiomyocytes
  - Endothelial cell
  - Epithelial cell
  - Fibroblast
  - hiPSC-CM
  - hiPSC-FB
  - Immune cell
  - Leukocytes
  - Lymphocytes
  - Macrophage
  - Monocytes
  - Natural killer cells
  - Neuronal cell
  - Neurones
  - Neutrophils
  - Oocyte
  - Pericytes
  - Platelet
  - Smooth muscle cells
  - T cells
- Isolated cells
  - Adipocyte
  - Cardiomyocyte
  - Dendritic cell
  - Embryonic cardiomyocytes
  - Endothelial cell
  - Epithelial cell
  - Fibroblast
  - hiPSC-CM
  - hiPSC-FB
  - Immune cell
  - Leukocytes
  - Lymphocytes

- Macrophage
- Monocytes
- Natural killer cells
- Neuronal cell
- Neurones
- Neutrophils
- Oocyte
- Pericytes
- Platelet
- Smooth muscle cells
- T cells

- Tissue chunk
- Tissue section (thin)
- Tissue slice
- Whole organ

**Oncogenes**

- cKIT-D816
- KRAS-G12D
- FLT3-ITD
- NPM ALK
- MLL-AF9
- IDH1
- VHL
- ATF6
- BRAF-V600E
- CTNNB1
- PTEN loss / KMT9A
- BAP1
- KRAS
- KMT2A
- AKT1
- cMyc
- NFE2

**Sample Processing**

- Cleared fixed tissue
- Formaldehyde-fixed and paraffin-embedded (FFPE)
- Formaldehyde fixation
- High-pressure frozen
- None (Physiological solution)
- OCT embedded and frozen

**Readout**

- Biomechanics
    - Cell stretching
    - Nanoindentation
    - Sarcomer Length
    - Single cell stretching
    - Tissue stretching
    - Tissue stretching Myodish
- Clinical Imaging
    - Computed tomography scan
    - Echocardiography
    - Magnetic resonance imaging

| | |
|---|---|
| | ◦ Positron emission tomography<br>• Computational modeling<br>• Electron microscopy<br>• Electrophysiology (clinical)<br>◦ Patch Clamp - current clamp<br>◦ Patch Clamp - voltage clamp<br>◦ Sharp Electrode<br>• Electrophysiology (lab)<br>• Flow cytometry / FACS<br>• Light microscopy<br>◦ Confocal<br>◦ Multiphoton<br>◦ Slidescanner<br>◦ Widefield<br>• Molecular Biology<br>◦ Aptamer (Proteomics)<br>◦ Bulk chromatin accessibility<br>◦ DNA Methylation (array)<br>◦ Enzyme-linked immunosorbent assay (ELISA)<br>◦ Gene Expression (array)<br>◦ Genotyping array and imputation<br>◦ Proximity extension assay (PEA)<br>◦ Quantitative polymerase chain reaction (qPCR)<br>◦ Standard biochemical assay<br>◦ Westernblot<br>• Optical mapping<br>• Optoacoustics<br>• Sequencing<br>◦ ATAC sequencing<br>◦ DNA methylation (WGBS)<br>◦ RNA sequencing<br>◦ Single Cell chromatin accessibility<br>◦ Single cell RNA seq<br>◦ Whole exome sequencing<br>◦ Whole genome sequencing<br>• Mass Spectrometry<br>◦ Interaction proteomics<br>◦ Metabolomics<br>◦ Proteomics<br><br>Complete this step entirely before proceeding to the next step. |
| **4** | **Consolidation of Results**<br>Combine the results of Steps 2 and 3:<br>1. For each subheading, provide a comprehensive, comma-separated list of aspects identified in Step 3.<br>2. Include any biomedical entities identified in Step 2 that are listed in PubTator but were not included as part of the aspects from Step 3. |

**PubTator3.0 Application Link:**

https://chatgpt.com/g/g-67516c0cff048191912549e36ee550eb-pubtatorsearch

### Forest Plots – Proportions of correctly annotated entities

*Figure S1: Precision of schema-related annotation predictions for each paper (without supplement) considered in the study together with pooled precision, heterogeneity and $\tau^2$.*

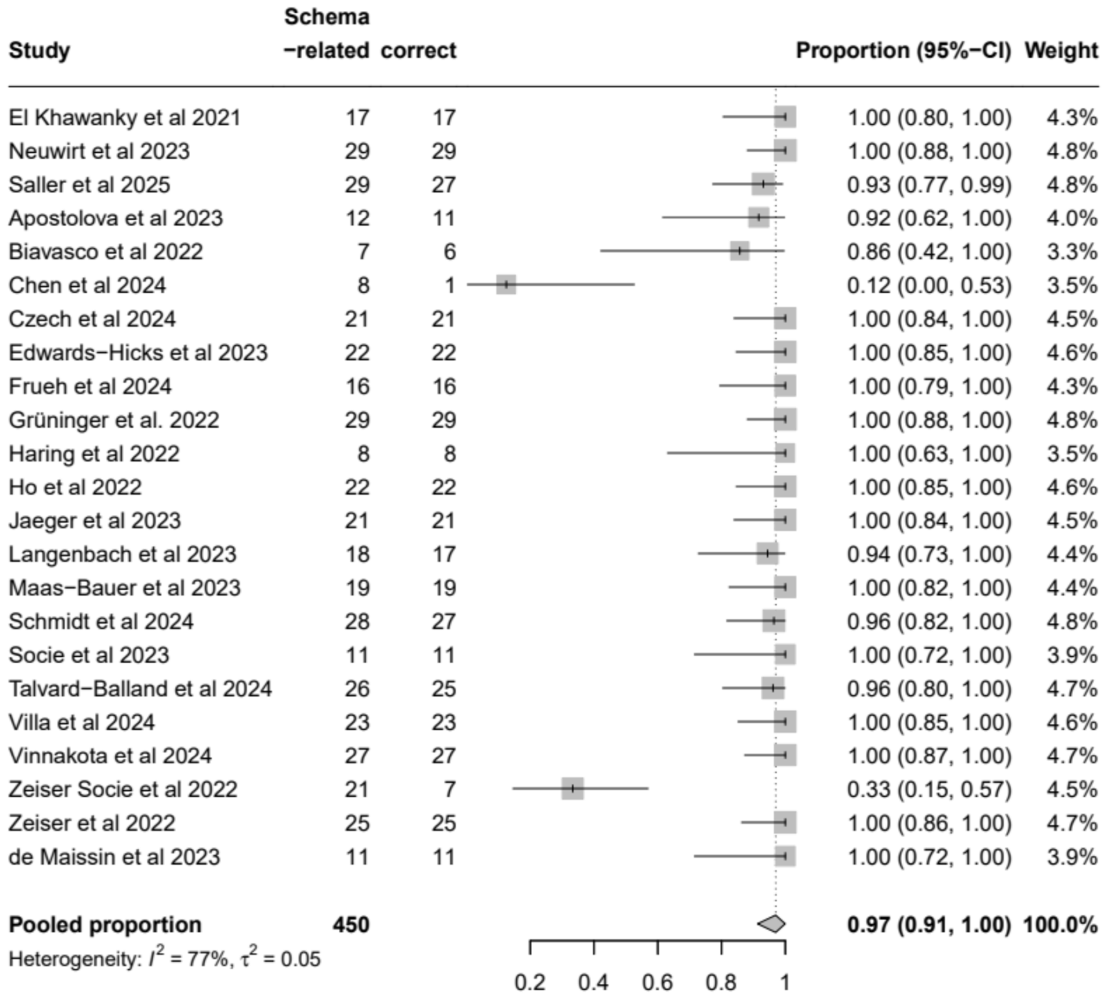

*Figure S2: Precision of PubTator-related annotation predictions for each paper (without supplement) considered in the study together with pooled precision, heterogeneity and τ².*

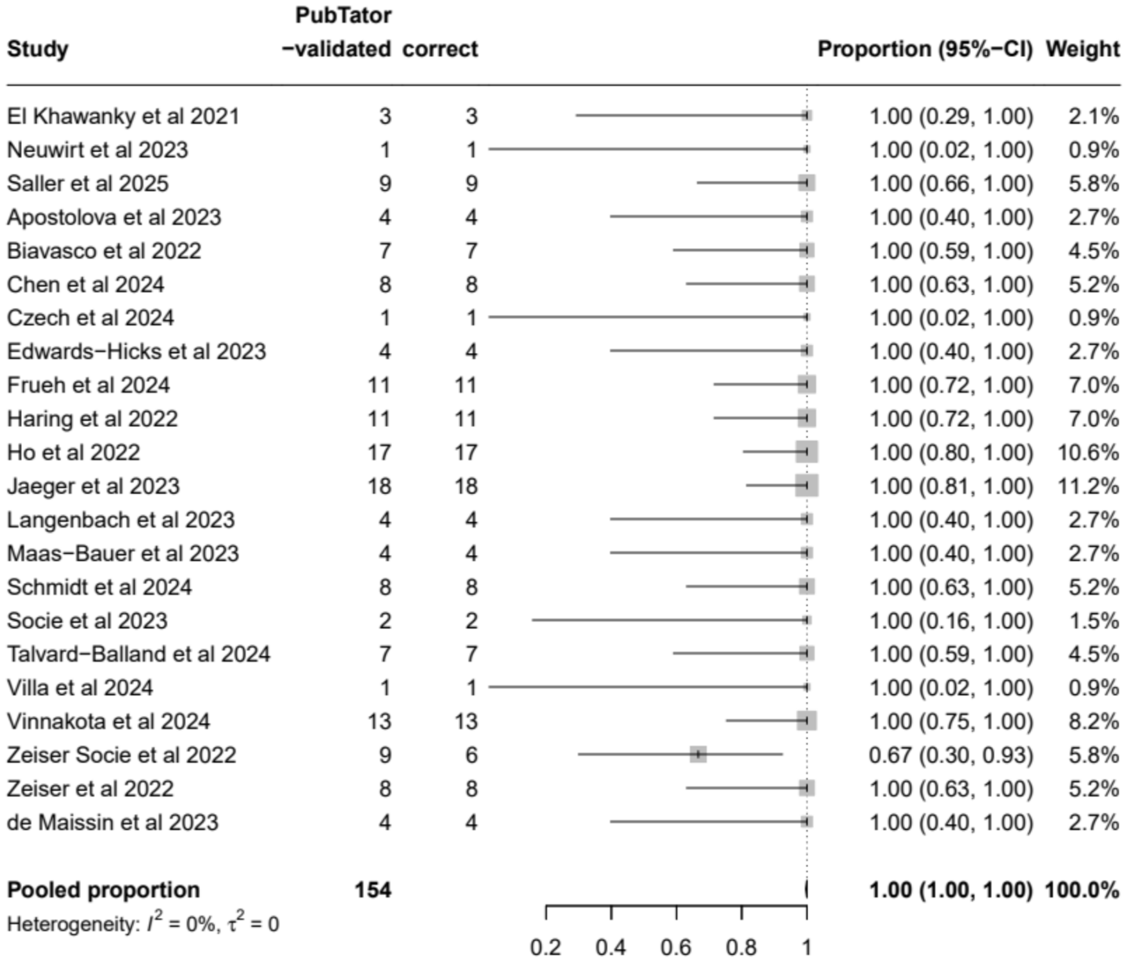

*Figure S3: Precision of PubTator-related annotation predictions for each paper with supplement (N=20) considered in the study together with pooled precision, heterogeneity and $\tau^2$.*

| Study | Schema −related | PubTator −validated | Total | correct | Proportion (95%−CI) | Weight |
|---|---|---|---|---|---|---|
| Saller et al 2025 | 16 | 1 | 17 | 14 | 0.82 (0.57, 0.96) | 4.4% |
| Apostolova et al 2023 | 14 | 5 | 19 | 19 | 1.00 (0.82, 1.00) | 4.6% |
| Biavasco et al 2022 | 10 | 13 | 23 | 22 | 0.96 (0.78, 1.00) | 4.9% |
| Chen et al 2024 | 18 | 6 | 24 | 11 | 0.46 (0.26, 0.67) | 4.9% |
| Czech et al 2024 | 21 | 1 | 22 | 22 | 1.00 (0.85, 1.00) | 4.8% |
| Edwards−Hicks et al 2023 | 15 | 7 | 22 | 22 | 1.00 (0.85, 1.00) | 4.8% |
| Frueh et al 2024 | 26 | 1 | 27 | 27 | 1.00 (0.87, 1.00) | 5.1% |
| Grüninger et al. 2022 | 23 | 12 | 35 | 35 | 1.00 (0.90, 1.00) | 5.5% |
| Haring et al 2022 | 28 | 10 | 38 | 37 | 0.97 (0.86, 1.00) | 5.6% |
| Ho et al 2022 | 29 | 9 | 38 | 38 | 1.00 (0.91, 1.00) | 5.6% |
| Jaeger et al 2023 | 15 | 11 | 26 | 26 | 1.00 (0.87, 1.00) | 5.1% |
| Langenbach et al 2023 | 21 | 2 | 23 | 23 | 1.00 (0.85, 1.00) | 4.9% |
| Maas−Bauer et al 2023 | 23 | 5 | 28 | 28 | 1.00 (0.88, 1.00) | 5.2% |
| Schmidt et al 2024 | 34 | 9 | 43 | 42 | 0.98 (0.88, 1.00) | 5.8% |
| Socie et al 2023 | 11 | 11 | 22 | 22 | 1.00 (0.85, 1.00) | 4.8% |
| Talvard−Balland et al 2024 | 30 | 12 | 42 | 41 | 0.98 (0.87, 1.00) | 5.7% |
| Villa et al 2024 | 29 | 3 | 32 | 32 | 1.00 (0.89, 1.00) | 5.4% |
| Vinnakota et al 2024 | 19 | 7 | 26 | 26 | 1.00 (0.87, 1.00) | 5.1% |
| Zeiser Socie et al 2022 | 8 | 5 | 13 | 12 | 0.92 (0.64, 1.00) | 3.9% |
| de Maissin et al 2023 | 11 | 3 | 14 | 13 | 0.93 (0.66, 1.00) | 4.0% |
| **Pooled proportion** | | | **534** | | **0.98 (0.95, 1.00)** | **100.0%** |

Heterogeneity: $I^2 = 67\%$, $\tau^2 = 0.02$

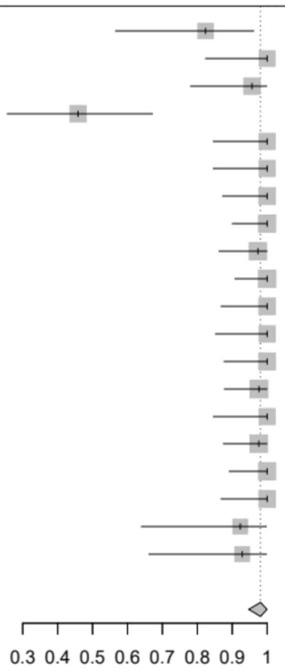

**Counts of annotation suggestions**

Additional distributions to show compatibility of number of annotation suggestions when the paper supplement is considered or not (Figure S4), as well as number of counts of annotation suggestions for the papers where no supplemental material was considered (Figure S5).

*Figure S4: Difference in counts of annotation suggestions for the papers for which the supplement was (1) not considered (2) considered (N=20).*

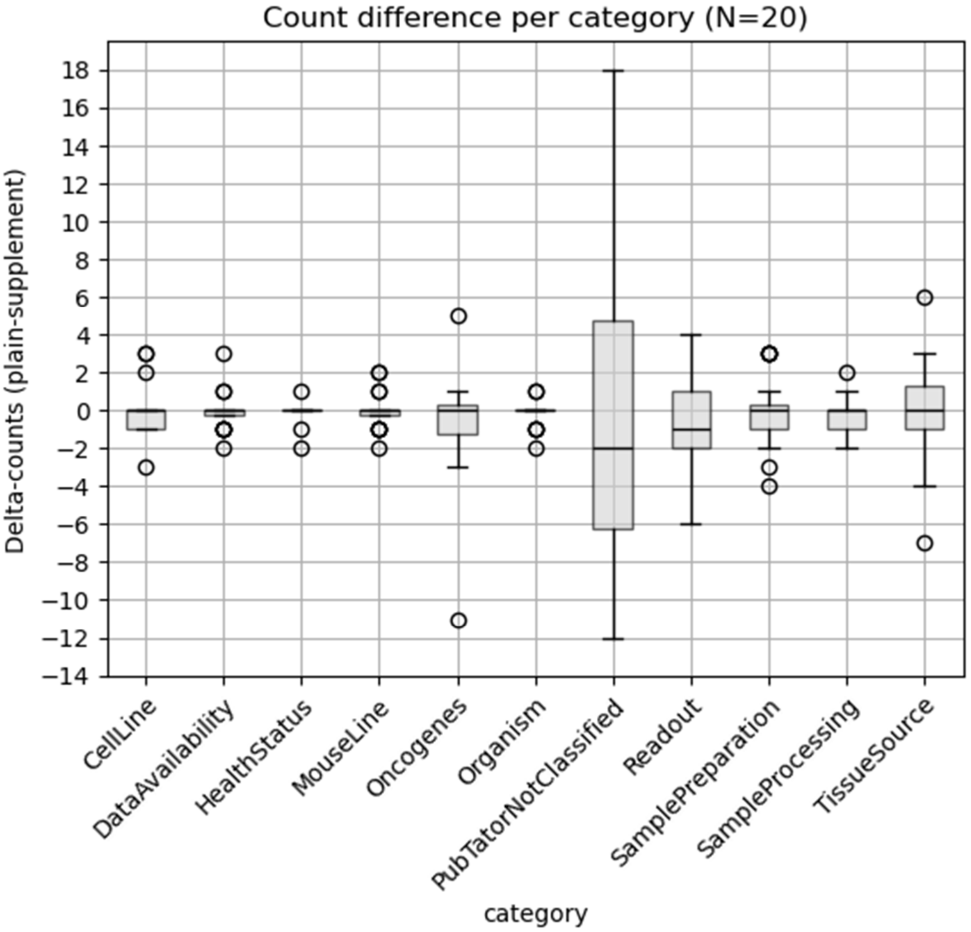

*Figure S5: Count spread of LLM annotation suggestions per category for papers with supplement not considered (N=23).*

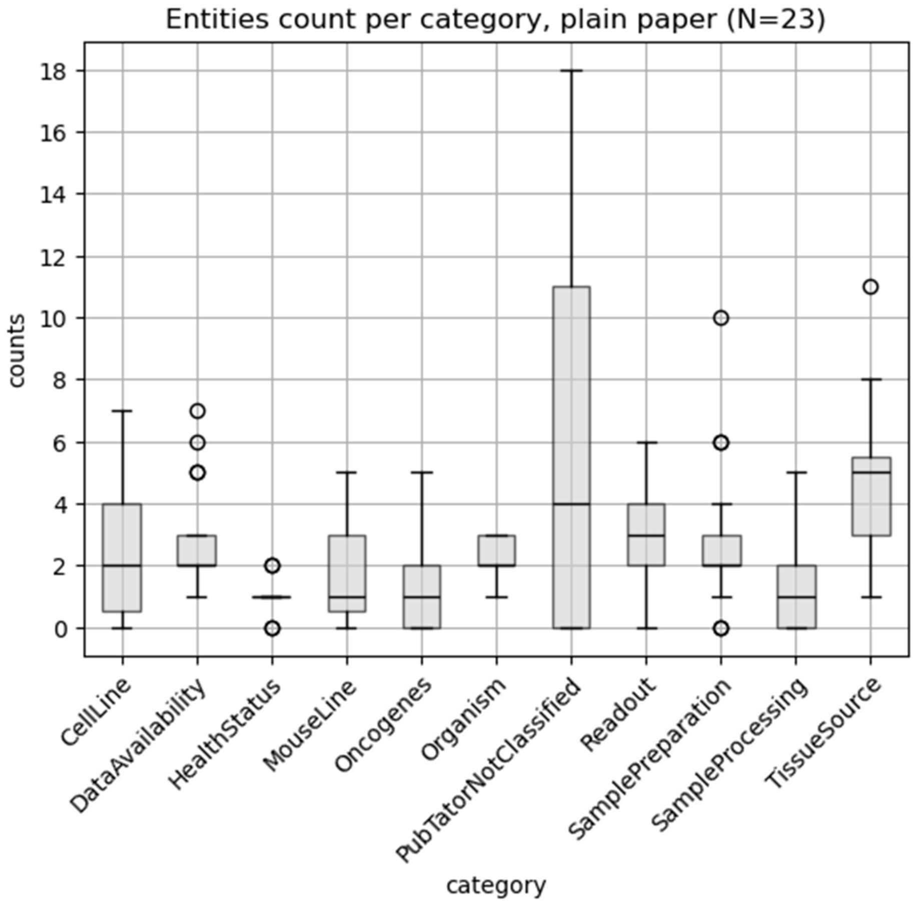


**Wilcoxon Signed-Rank-Test Table**

*Table S3: Wilcoxon test results (p-value) for compatibility of number of suggested entries between paper without and with supplement (N=20), per category and overall number of suggestions (without Data availability).*

| category | p-value |
|---|---:|
| CellLine | 0.794 |
| DataAvailability | 0.660 |
| HealthStatus | 0.414 |
| MouseLine | 0.951 |
| Oncogenes | 0.265 |
| Organism | 0.317 |
| PubTatorNotClassified | 0.706 |
| Readout | 0.365 |
| SamplePreparation | 0.750 |
| SampleProcessing | 0.222 |
| TissueSource | 1.000 |
| **Overall result** | **0.556** |
